# Analysis of plasma DNA fragmentation patterns from dried blood spots

**DOI:** 10.1101/2024.12.22.24319522

**Authors:** Michelle D. Stephens, Elise C. Dietmann, Bradon R. McDonald, Everlyne N. Nkadori, Syed Nabeel Zafar, Stephanie M. McGregor, Muhammed Murtaza

## Abstract

Circulating tumor DNA analysis holds promise for early detection of cancer. However, costs and complexity of genomic methods, and logistical requirements for blood collection and processing limit large scale implementation. Here, we evaluated the feasibility of genome-wide analysis of fragmentation patterns in plasma DNA obtained from dried blood spots (DBS). Across 75 individuals including 25 patients with cancer, we prepared conventional DBS, and plasma-separating DBS (psDBS) from whole blood samples, prior to plasma separation. In paired comparisons of plasma DNA between psDBS and blood tubes, we observed correlated concentrations, similar distributions of fragment lengths, correlated genome-wide fragmentation features, and equivalent performance of machine learning algorithms for cancer detection. Our results provide proof-of-principle that plasma DNA fragmentation analysis can be performed from small amounts of DNA obtained from psDBS. With further development and validation, this approach can expand the reach and impact of blood-based early cancer detection, particularly for resource constrained environments such as low- and middle-income countries and rural healthcare systems.

## Main Text

Circulating tumor DNA analysis has shown promising results for cancer detection and response monitoring during treatment(*1*). Early work in this field relied on deep molecular analysis of a few genomic loci with known or recurrent somatic genomic alterations and methylation changes(*2–4*). More recently, new approaches have traded high sequencing depth at a few loci with wide breadth of genomic coverage at low depth(*5, 6*). Low-depth whole genome sequencing methods analyze variation in genome-wide features such as sequencing coverage (driven by copy number alterations), mutation signatures, fragmentomic features, and nucleotide frequencies surrounding fragment ends(*5–9*). Incorporating several thousand or more loci from across the genome into these features enables high accuracy while reducing requirements of sequencing depth, amount of input DNA for sequencing library preparation, and sequencing costs(*10*). However, unlike somatic single nucleotide variants, differences in fragmentation features are not inherently tumor specific, making them more susceptible to pre-analytical sources of variation(*11–13*). Wide adoption of plasma DNA fragmentation analysis for cancer detection and monitoring is hindered by stringent upstream requirements for collection, processing, and storage of blood samples. In resource-limited environments, particularly low- and middle-income countries (LMICs), the costs associated with phlebotomy, sample processing to isolate plasma, storage, and shipping while maintaining an effective cold chain can be prohibitive(*14*). These challenges reduce the likelihood that patients will receive molecular testing and perpetuate disparities in cancer detection and outcomes(*15*). An inexpensive, and logistically simpler alternative has immense potential to bring blood-based early detection of cancer to resource limited environments.

We have recently shown that robust fragmentation analysis is feasible from sequencing data representing 1 to 10 million plasma DNA fragments from each sample (equivalent to genomic coverage of 0.05 to 0.5x)(*5*). At the reported mean concentration of 4 ng/mL or 1200 haploid genome equivalents/mL of plasma DNA in healthy individuals, adequate amount of plasma DNA for shallow whole genome sequencing should be available in as little as 100 to 150 µL of blood (3-5 drops of blood). Here, we test this hypothesis and evaluate whether plasma DNA fragmentation patterns can be analyzed from a few drops of blood, when collected as dried blood spots (DBS).

From a total of 75 participants including 45 healthy individuals, 25 patients with cancer and 5 patients with non-malignant disease, we collected blood samples and spotted conventional DBS (cDBS) as well as plasma-separating DBS (psDBS) from whole blood prior to further processing and isolation of plasma (Fig. 1 and table S1). All paired samples underwent DNA extraction and were analyzed using shallow whole genome sequencing (WGS) performed without any further DNA fragmentation. Mean coverage was 0.27x for plasma DNA from blood tubes (SD 0.11), 0.24× for cDBS (SD 0.11), and 0.14× for psDBS (SD 0.07).

**Fig. 1.**
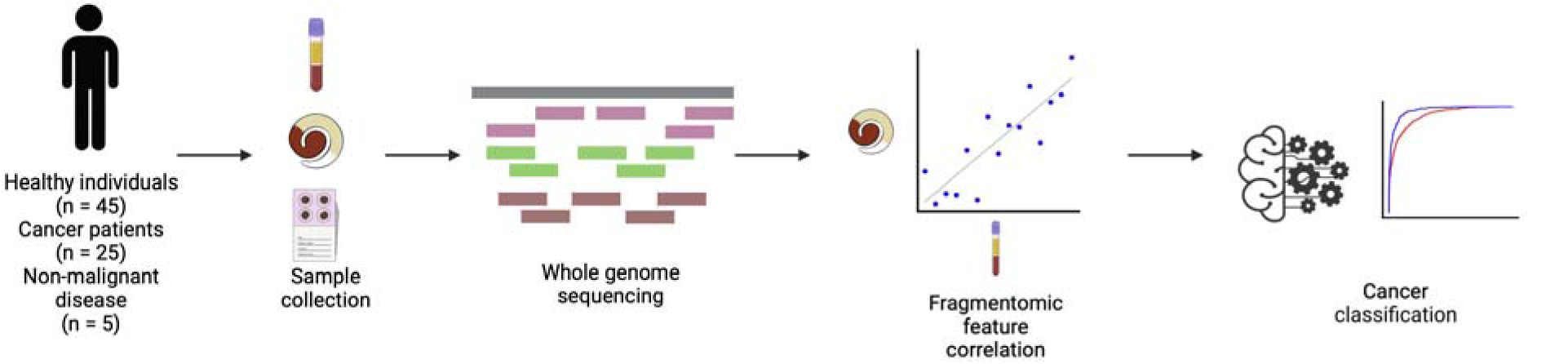
Study overview. We collected matched plasma samples from blood tubes, plasma-separating dried blood spots, and conventional dried blood spots from 75 individuals. Samples from 45 healthy individuals, 25 patients with cancer, and 5 patients with non-malignant conditions including pancreatic cysts and pelvic mass were included in this study (table S2). After DNA extraction from each sample type, we performed whole genome sequencing and measured multiple features related to plasma DNA fragmentation. We compared these features between paired samples to assess similarity and correlation. We also compared performance of machine learning classifiers built to distinguish cancer from healthy samples, across paired sample sets.

### Evaluation of plasma DNA concentration and fragment lengths

Following DNA extraction and elution, plasma DNA concentration was too low in DBS samples for accurate quantification using fluorometric or PCR-based methods. However, for one set of outlier samples where plasma DNA concentration in the blood tube was 127 ng/ml, electrophoresis of extracted DNA showed a fragment length distribution consistent with plasma cell-free DNA in psDBS and a large fraction of DNA >1000 bp in cDBS (fig. S1). In addition, whole genome libraries prepared using extracted DNA from psDBS and cDBS showed length distributions consistent with plasma DNA libraries prepared from blood tubes (fig. S2). To infer whether DNA concentration was preserved in DBS, we measured DNA concentration in plasma from blood tubes and compared it with the yield of WGS libraries from each set of DBS samples (such that all compared samples underwent the same number of amplification cycles). Library yield was correlated with plasma DNA concentration in blood tubes when WGS libraries were prepared from psDBS (r^2^=0.46; n=74; p=2.6 x 10^-11^, Fig. 2A), but not when libraries were prepared from cDBS (r^2^=0.14, n=43, p=1.4 x 10^-2^; Fig. 2B). One sample with very high plasma DNA concentration in the blood tube (127 ng/ml) was excluded from this analysis.

**Fig 2.**
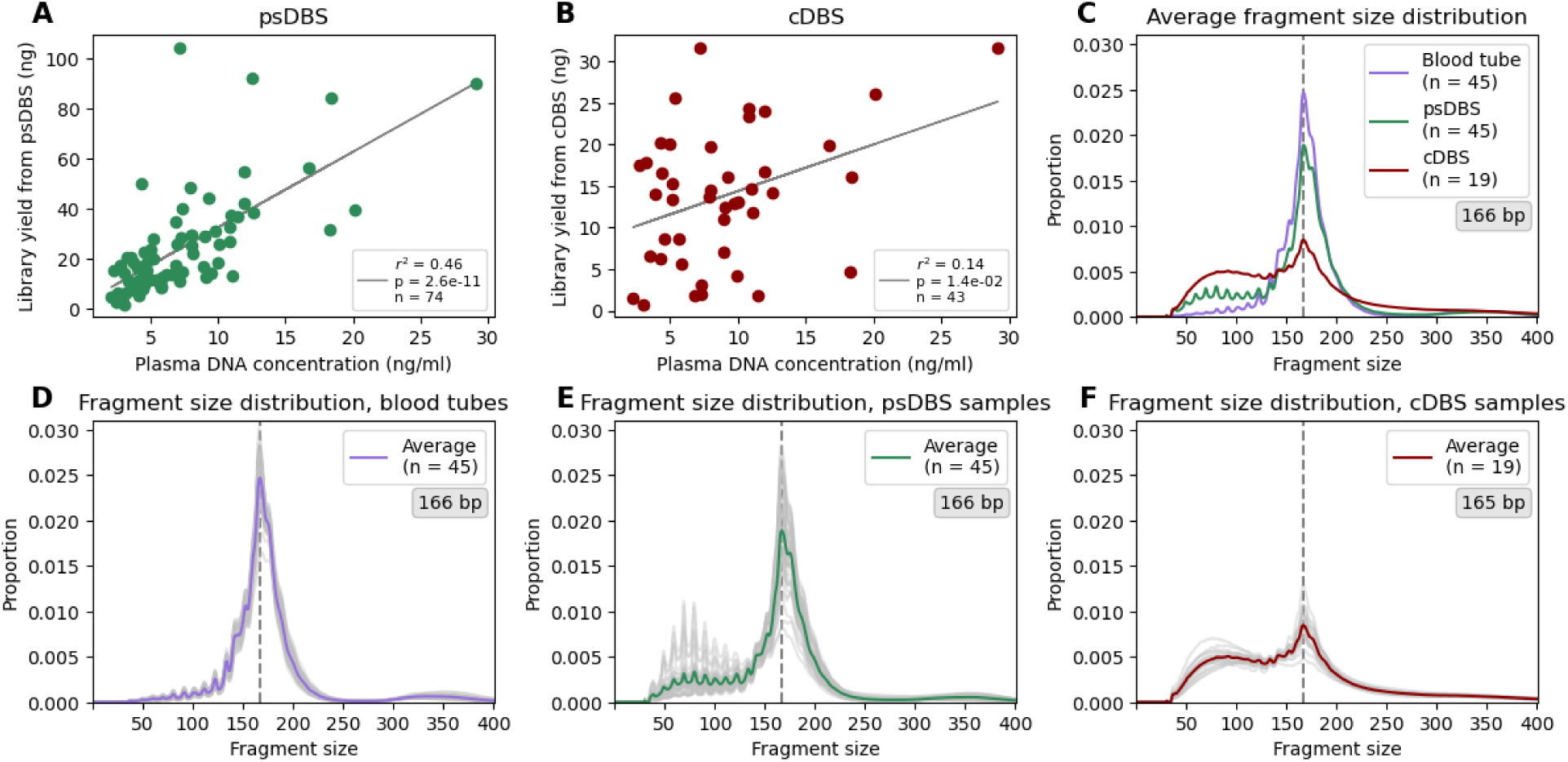
Comparison of plasma DNA whole genome sequencing libraries prepared from blood tubes, plasma-separating dried blood spots and conventional dried blood spots. **A.** Comparison of plasma DNA concentration in blood tubes with library yield from matched psDBS (one outlier is excluded in this analysis, with a plasma concentration of 127.2 ng/ml and a psDBS library yield of 450 ng) **B.** Comparison of plasma DNA concentration in blood tubes with library yield from matched cDBS (one outlier is excluded in this analysis, with a plasma concentration of 127.2 ng/ml and cDBS library yield of 87.6 ng) **C.** Average fragment size distribution observed in plasma DNA whole genome libraries from blood tubes, psDBS, and cDBS from healthy individuals. **D.** Fragment size distribution of plasma DNA whole genome libraries from blood tubes from healthy individuals. Each individual sample is one grey line, with the average plotted in purple. **E.** Fragment size distribution of whole genome libraries from psDBS samples from healthy individuals. Each individual sample is one grey line, with the average plotted in green. **F.** Fragment size distribution of whole genome libraries from cDBS samples from healthy individuals. Each individual sample is one grey line, with the average plotted in red.

To evaluate whether plasma DNA fragment lengths were preserved in DBS, we compared insert size distributions measured using WGS data in samples obtained from healthy individuals. The modal size of all three sample sets was 165 – 166 bp, similar to prior reports of plasma DNA fragment lengths associated with mono-nucleosomes (Fig. 2C)(*16*). Plasma DNA libraries from blood tubes and psDBS showed a similar and large proportion of fragments at this size, compared to cDBS samples which showed a smaller proportion of mono-nucleosome length fragments. Blood tubes and psDBS samples also showed a 10-bp periodicity below 130 bp, suggestive of enzymatic degradation of nucleosome-associated DNA as previously described(*7*). In addition, both sample types showed very few fragments between 230 and 300 bp, and a small second modal peak at approximately 340 bp, a fragment length associated with DNA preserved in di-nucleosomes. In comparison, cDBS libraries showed a far higher proportion of fragments between 40 and 150 bp without 10 bp periodicity and did not show a distinct peak of di-nucleosomal length. For plasma samples from blood tubes, fragment length distributions were highly conserved across samples (Fig. 2D). For psDBS samples, most samples had a similar distribution, but a subset had very high contribution of fragments under 120 bp, displaying strong periodicity (Fig. 2E). For cDBS samples, fragment size distributions were highly variable (Fig. 2F).

### Evaluation of genome-wide plasma DNA fragmentation features

To evaluate whether genome-wide plasma DNA fragmentation patterns were preserved in DBS, we measured multiple fragmentation features using WGS and compared them across the paired sample sets.

In recent work, we have developed a cancer detection assay called genome-wide analysis of fragment ends (GALYFRE), that incorporates multiple fragmentomic features measured using WGS into a machine learning model(*5*). One feature included in GALYFRE is aberrant fragmentation score (AFS), a measurement based on the fraction of fragment ends observed within nucleosome-associated recurrently protected genomic regions in each sample. In the current study, we found no correlation in AFS between blood tubes and cDBS (Fig. 3A, r^2^=0.0, n=44, p=0.92), but observed a positive correlation between blood tubes and psDBS (Fig. 3B, r^2^=0.24, n=75, p=9.5 x 10^-6^). We observed that in a subset of psDBS where the AFS was higher than the corresponding plasma samples from blood tubes, the psDBS samples also had disproportionate contributions of fragments <=120 bp. Once psDBS samples with >20% of fragments <=120 bp were excluded (n=17, 23% of total samples), the coefficient of determination (r^2^) for the linear relationship between blood tubes and psDBS samples improved to 0.66 (n=58, p=5.6 x 10^-16^; Fig. 3C). Across all three sample sets, AFS in cDBS samples were much higher while they remained comparable between blood tubes and psDBS.

**Fig 3.**
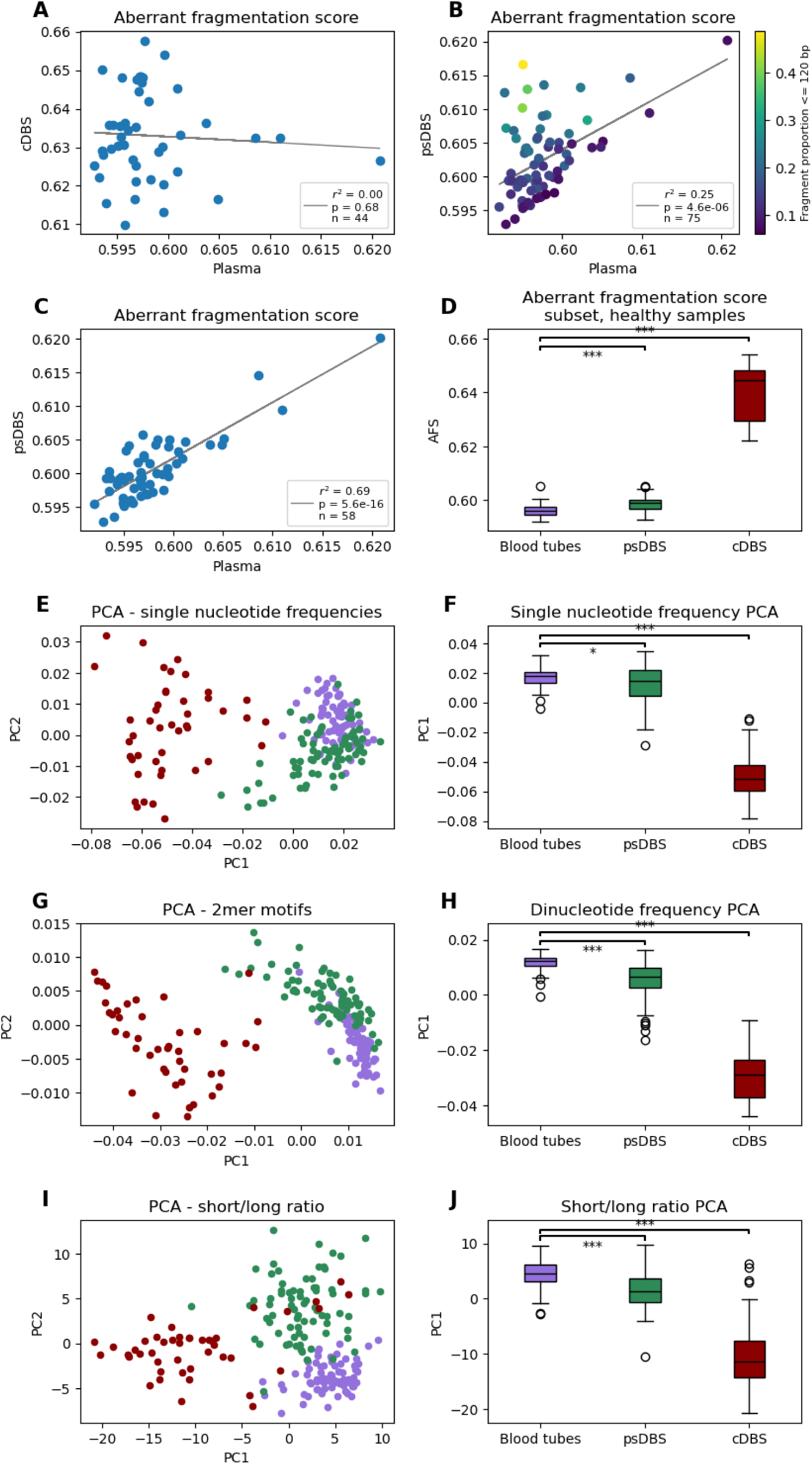
Comparison of plasma DNA fragmentation features between matched blood tubes, plasma-separating dried blood spots and conventional dried blood spots. **A.** Correlation in aberrant fragmentation score (AFS) between matched blood tubes and cDBS samples. **B.** Correlation in AFS between matched blood tubes and psDBS samples. The color scale indicates proportion of fragments in the psDBS sample of less than or equal to 120 bp. **C.** Correlation in AFS between matched blood tubes and psDBS samples, excluding samples with short fragment proportion of greater than 20%. **D.** Boxplot of AFS in healthy samples, excluding matched samples with a psDBS short fragment proportion of greater than 20%. **E.** Principal component analysis for nine selected single nucleotide frequencies from 5’ fragment ends, previously described in a cancer detection machine learning model. **F.** Box plot comparing principal component 1 for single nucleotide frequencies from 5’ fragment ends across all three sample types. **G.** Principal component analysis for dinucleotide frequencies from 5’ fragment ends. **H.** Box plot comparing principal component 1 for dinucleotide frequencies from 5’ fragment ends across all three sample types. **I.** Principal component analysis for the ratio of short to long fragments in 544 bins of 5000 kbp each from across the genome. **J.** Box plot comparing principal component 1 for ratio of short to long fragments across all three sample types.

Another set of fragmentomic features included in GALYFRE was nucleotide frequencies surrounding fragment ends(*5*). In the current study, we compared mean fragment end nucleotide frequencies, measured for each position surrounding the 5’end of plasma DNA fragments, averaged across all fragments in a sample. Using principal components to analyze single nucleotide frequencies at plasma DNA fragment ends, we found that blood tubes overlapped with psDBS samples, but cDBS samples were clearly separated in the first principal component (Fig. 3EF). Individual nucleotide frequencies at each position surrounding the 5’end of DNA fragments showed better correlation between psDBS and blood tubes than between cDBS and blood tubes (fig. S3 and S4). Similarly, when considering dinucleotide frequencies at the 5’ fragment end(*9, 17*), we found that blood tubes overlapped together with psDBS, while cDBS was separated from the two in the first principal component (Fig. 3GH). Using hierarchical clustering based on dinucleotide frequencies, psDBS and plasma samples from blood tubes clustered together while cDBS showed clear differences and clustered separately (fig. S5). The most common dinucleotide observed at the 5’ fragment ends in blood tubes and psDBS was CC. Using a similar approach to evaluate 4-mer motifs at fragment ends, we found psDBS and plasma from blood tubes were more similar to each other, compared to cDBS (fig. S6 and fig. S7).

One recent study showed that genome-wide variation in the ratio of short fragments (100 to 150 bp) to long fragments (151 to 220 bp)(*6*) can be informative for cancer detection. Here, we measured the ratio of short to long fragments for 544 bins of 5000 kbp across the genome. Using principal components analysis, we found that blood tubes and psDBS samples overlapped in the first principal component while cDBS samples showed a clear difference (Fig. 3IJ).

### Classification between cancer and healthy individuals using plasma DNA fragmentation analysis

Encouraged by results from comparisons of fragmentation features, we evaluated the potential to detect cancer-derived DNA using psDBS samples compared to corresponding plasma samples from blood tubes. Blood tube and psDBS samples were collected from 25 patients with cancer across multiple cancer types (breast, head and neck, pancreas, endometrium, kidney, prostate, and gall bladder). 21/25 samples (84%) were collected from patients with Stage I-III cancer (table S2). We first evaluated whether copy number aberrations were detectable in plasma DNA from blood tubes. Of 25 patients with cancer, we found copy number aberrations detectable in plasma from 2 patients. Copy number aberrations detected in corresponding psDBS samples were nearly identical (Fig. 4A and fig. S8).

**Fig 4.**
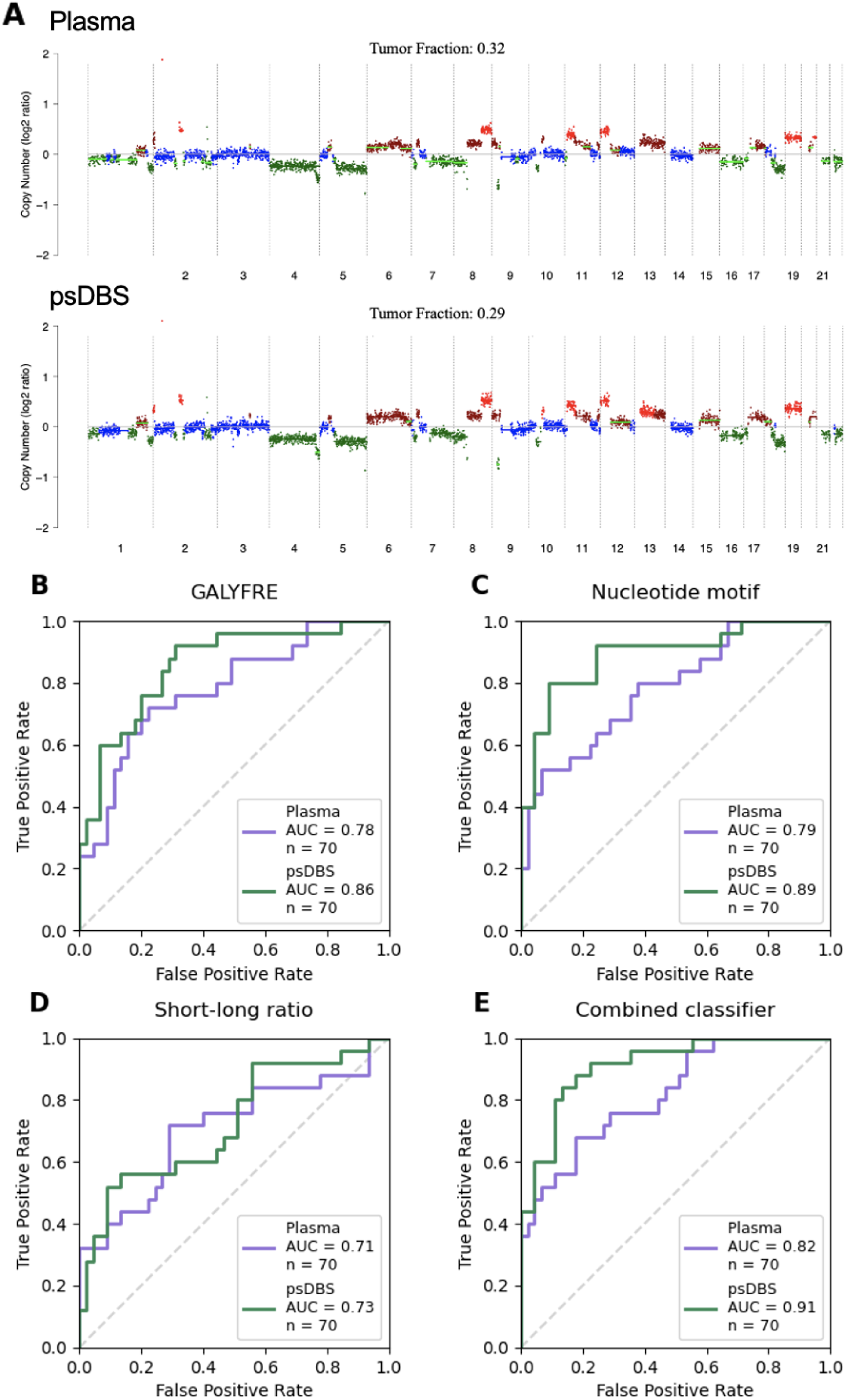
Comparison of plasma DNA between healthy individuals and patients with cancer using blood tubes and psDBS. **A.** Comparison of copy-number aberrations and inferred tumor fraction in plasma DNA between matched blood tubes (top) and psDBS (bottom) from a patient with Stage IV pancreatic cancer. **B.** ROC curves evaluating accuracy of the GALYFRE classifier based on 10 fragmentomic features including aberrant fragmentation scores and frequencies of 9 single nucleotides at loci surrounding 5’ fragment ends, using blood tubes (purple) and psDBS (green). **C.** ROC curves evaluating accuracy of a random forest machine learning model based on 4-mer nucleotide motifs at 5’ fragment ends, using blood tubes (purple) and psDBS (green). **D.** ROC curves evaluating accuracy of a random forest machine learning model based on the ratio between short and long plasma DNA fragments in 544 bins of 5000 kbp each across the genome, using blood tubes (purple) and psDBS (green). **E.** ROC curves evaluating accuracy of an ensemble model that combines the GALYFRE classifier (B), and models based on nucleotide motifs (C) and short-to-long fragment ratios (D), using blood tubes (purple) and psDBS (green).

Next, we evaluated performance of multiple machine learning models based on different sets of fragmentation features to classify between cancer patients and healthy individuals. For each model, we compared its performance using plasma DNA WGS from blood tubes and corresponding psDBS samples. Using GALYFRE, a random forest model that incorporates AFS and nucleotide frequencies surrounding fragment ends(*5*), we observed an area under the receiver operating characteristic curve (AUROC) of 0.78 and 0.86 for blood tubes and psDBS samples, respectively (Fig. 4B, p=0.27). We also evaluated the performance of a random forest model based on the frequency of 256 4-mer motifs observed at the 5’end of DNA fragment ends, as recently described(*9*). We observed an AUROC of 0.79 and 0.89 for blood tubes and psDBS samples, respectively (Fig. 4C, p=0.11). We further evaluated the performance of a random forest model incorporating short-to-long DNA fragment length ratios measured in each of 544 bins of 5000 kbp across the genome, as recently described(*6*). We observed an AUROC of 0.71 and 0.73 for blood tubes and psDBS samples, respectively (Fig. 4D, p=0.84). Finally, we built an ensemble model incorporating all these features together. Using this approach, we observed an AUROC of 0.82 and 0.91 for blood tubes and psDBS samples, respectively (Fig. 4E, p=0.06).

## Discussion

Detection and treatment of cancer at earlier stages of disease dramatically increases the likelihood of achieving cure. Several studies have demonstrated the relevance of circulating tumor DNA analysis for cancer detection, either as a multicancer test(*4*) or for individual cancer types(*18, 19*). Methods for circulating tumor DNA analysis have evolved from targeted molecular assays to genome-wide approaches that integrate hundreds to thousands of genomic loci to generate features for machine learning models and achieve high classification accuracy(*11*). Such genome-wide approaches often rely on shallow sequencing coverage and require limited amounts of input DNA. In earlier work, we had performed in silico analysis to show that plasma DNA fragmentation patterns could be reliably measured using data representing only 1-2 million fragments(*5*). Here, we have tested this hypothesis and found that plasma DNA fragmentation patterns are largely preserved in plasma-separating dried blood spots. In addition, we found that psDBS can achieve equivalent performance for cancer detection when compared with corresponding, paired plasma samples obtained from blood tubes.

Since concentration of DNA in plasma is generally much lower than the peripheral blood cells compartment, the field of circulating tumor DNA analysis has long insisted on stringent conditions for sample processing and storage to ensure accurate results(*13, 20*). For large multisite clinical trials, this increases costs associated with sample collection for ctDNA analysis because it requires either trained local biospecimen processing teams at each site or real-time shipping of samples collected in specialized cell-free DNA blood tubes to a central biorepository. Recent studies showed that cDBS can potentially overcome these limitations for monitoring of treatment response in patients with metastatic cancer using copy number analysis, but this requires removal of high molecular weight background DNA fragments shed by peripheral blood cells (*21, 22*). Here, we have found that when plasma is separated from the blood cell fraction at the time of blood spot collection, plasma DNA fragmentation patterns are preserved in psDBS and can generate insights for early cancer detection that are similar to corresponding blood tubes.

Collection of psDBS is straightforward and could be performed in ambulatory care or self-collected at home, without requiring a trained phlebotomist. Once the blood spot has dried, there is no additional processing or cold-chain storage required, and blood spots can be shipped at ambient temperature to a central lab for processing(*23*). These improvements in logistics of sample collection can enable future clinical studies and applications such as early detection of cancer in primary care patients and distributed monitoring of treatment response. In addition, this approach can enable the expansion of current advances in cancer detection and monitoring to underserved populations in resource-limited settings, including LMICs and rural healthcare systems. Addressing this challenge is urgent and critical, since the burden of cancer is rapidly rising in LMICs where nearly two-thirds of all new cancer cases and 70% of all cancer deaths are projected to occur by 2030 (*24*).

While our results hold tremendous potential to increase access to blood-based early detection methods, there are limitations of this study that should be considered. First, to enable evaluation of multiple blood spots from each participant and comparison with blood tubes obtained at the same time, we collected blood tubes from patients and then prepared cDBS and psDBS from whole blood. This allowed us to minimize participant discomfort while evaluating different sample collection devices, and to establish the minimum volume of blood sample needed for DBS. As a result, our analysis of psDBS represents DNA fragmentation patterns measured from venous blood rather than capillary blood collected from fingersticks. One study using PCR amplification of a repetitive genomic locus using multiple amplicon sizes showed no difference in DNA integrity between venous and capillary cfDNA(*25*). In ongoing work, we are expanding our study to include psDBS samples directly collected by fingerstick. Second, we found that a subset of psDBS samples (∼20%) showed a high abundance of ultrashort fragments which are not observed in corresponding plasma samples. We observed no clear association between this phenomenon and any biological or clinical factors associated with the participant or sample, suggesting this may be arising from variation in device manufacturing or DNA extraction protocols. This is an area of further investigation. Third, we found that fragmentation patterns in psDBS are highly correlated with and similar to plasma from blood tubes but not completely equivalent. Hence, we expect that any existing computational models for analysis of plasma DNA fragmentation previously built using plasma from blood tubes will require re-training and validation using psDBS samples, prior to implementation.

In summary, we report proof-of-principle results that analysis of DNA fragmentation patterns from plasma-separating dried blood spots is feasible and can overcome the current logistical barriers that limit scalability of plasma DNA testing. Pending further validation in larger studies and clinical trials, this approach can vastly improve access to early cancer detection as well as other diagnostic applications of plasma DNA fragmentation analysis beyond oncology(*26*).

## Acknowledgments

The authors wish to thank Judy Wang, Will Suter, Hellen Nginga, Marissa Lehman, Samantha Probelsky, Misbah Zaeem, and the UW Carbone Cancer Center BioBank for sample collection and processing.

## Author contributions

Conceptualization: MDS, MM

Methodology: MDS, MM

Investigation: MDS, ECD, BRM, ENN, SMM

Visualization: MDS, ECD

Funding acquisition: MM

Project administration: MM

Supervision: MM

Writing – original draft: MDS, ECD, MM

Writing – review & editing: MDS, ECD, BRM, ENN, SNZ, SMM, MM

## Data and materials availability

Genome sequencing data reported here will deposited in dbGaP upon acceptance. Code reproducing the reported analysis will be deposited to zenodo upon acceptance.

## Supplementary Materials

Materials and Methods

Figs. S1 to S8

Tables S1 to S2

