## Supplementary Materials for "Analysis of plasma DNA fragmentation patterns from dried blood spots"

**The PDF file includes:**

Materials and Methods
Figs. S1 to S8
Tables S1 to S2
References

### Materials and Methods

Enrollment and sample collection

Healthy volunteers and cancer patients were enrolled at the Translational Genomics Research Institute and by the University of Wisconsin-Madison Biobank. Samples were collected under protocols approved by Western Institutional Review Board (IRB, protocol number 20181812) and by University of Wisconsin-Madison IRB (protocol number 2016-0934). Samples from cancer patients were collected prior to surgery. No patients had received neoadjuvant chemotherapy.

Sample processing

All plasma samples were collected in EDTA tubes and centrifuged twice at room temperature. Plasma aliquots were stored at -80° C prior to DNA extraction.

For conventional dried blood spots (cDBS) and plasma-separating dried blood spots (psDBS), whole blood was collected in EDTA or preservative-free tubes. All samples collected in preservative-free tubes were spotted within 20 minutes. Samples collected in EDTA tubes were spotted within 1 hour. For all spots, whole blood was pipetted onto the respective devices. cDBS were prepared on QIAcard FTA micro cards (Qiagen) from 125 µl whole blood; psDBS were prepared on the HemaSpot SE device from 125 – 150 µl whole blood. Spots were fully dried before storage at room temperature in a zippered plastic bag.

DNA extraction

Plasma DNA was extracted from 1 mL plasma with the MagMAX Cell-Free DNA Isolation Kit (ThermoFisher) according to manufacturer instructions, using the EDTA tube workflow.

For cDNA and psDBS, DNA was extracted with the MagMAX Cell-Free DNA Isolation Kit (ThermoFisher) according to a modified protocol. For the cDBS, one-half to the full spot was excised and cut into quarters. For the psDBS, the entirety of the plasma-containing portion of the membrane was excised and cut into pieces with a maximum length of approximately 10 millimeters. Seven hundred fifty microliters of phosphate buffered saline was added to the sample, and the sample was lysed with proteinase K and sodium dodecyl sulfate according to manufacturer instructions. The supernatant, excluding any portions of the membrane, was collected and added to the binding solution/beads mix. All remaining aspects of the extraction protocol were unchanged from manufacturer instructions.

Extracted DNA was assessed and quantified with the cfDNA assay on the TapeStation system (Agilent).

Library preparation & sequencing

Whole genome libraries were prepared from plasma, DWBS, and DPS DNA. For plasma DNA, input was limited to a maximum of 2 ng, and 10 cycles of amplification were performed. For DWBS and DPS DNA, a fixed input of 12 µl (of a total elution volume of 30 µl) of the sample was used, and 14 cycles of amplification were performed. All libraries were prepared using the ThruPLEX DNA-Seq HV kit (TakaraBio). Libraries were quantified using the Tapestation D1000 HS assay (Agilent). Sequencing was performed on the NextSeq 1000/2000 system

(Illumina) to generate 100 base pair paired-end reads. Demultiplexing, trimming, and FASTQ conversion was performed on-board using DRAGEN (version).

Data processing
All sequencing data was aligned to human genome build hg19 (p13_105) using BWA-MEM. Files were converted to BAM format using SAMtools, and duplicate reads were marked. Analysis was limited to non-duplicate reads marked as properly paired with a minimum mapping quality of 60. The fragment size of all aligned reads was calculated using bedtools(*1*).Fragments between 30 and 1000 base pairs were analyzed.

Copy number analysis
Copy number analysis was performed on aligned sequencing data with ichorCNA_U (*2*), a fork of ichorCNA(*3*). Only somatic chromosomes were analyzed, and a bin size of 500 kb was used.

Measurement of aberrant fragmentation
Aberrant fragmentation scores (AFS) were calculated as previously described (*4*). To calculate AFS, the same map of recurrently protected regions (RPRs) as published was used. Fragments fully spanning an RPR were considered non-aberrant, while fragments with one or more ends falling within an RPR were considered aberrant. AFS was calculated as a proportion of aberrant fragments to all fragments intersecting an RPR, with adjustments for fragment length and GC content as previously described. Only fragments between 140 and 220 bp were considered for calculation of AFS.

Single nucleotide frequencies
Nucleotide frequencies were calculated as previously described (*4*). For all fragments between 50 and 1000 base pairs, the reference sequence from 10 base pairs outside the fragment end to the first 10 base pairs of the fragment end was collected with Homertools (*5*). Average frequencies at all positions were calculated based on the reference sequence.

End motifs
Counts of four-mer and two-mer end motifs were calculated for the primary alignment of all non-duplicate fragments. To assess similarity between sample types, hierarchical clustering was performed with Seaborn(*6*), using `average` as the linkage method and `Euclidean` as the distance metric.

Short / long fragments
Counts of short (100 to 150 bp) and long (151 – 220 bp) fragments were measured in 5000 kb bins across the genome. From 590 initial bins, only bins with at least 2000 long fragments were included in analysis, resulting in 544 bins. The ratio of short to long fragments in each bin was calculated for these 544 bins, and then each sample was normalized to a mean bin fragment ratio of 0, with a standard deviation of 1.

Modeling

Classification performance was assessed using four machine learning models. For all models, an ensemble model of 100 random forest models was generated using 100 iterations of crossvalidation, and the score for each sample was taken as the average of all scores when the sample was in the validation split. For each iteration, the data is split into an 80/20 train/validation split, and a random forest model is generated with a maximum depth of 5 and a minimum samples per leaf of 5.

The GALYFRE model was implemented with features and hyperparameters as described previously (*4*). Briefly, 10 features are used, including AFS, and 9 nucleotide frequency features at positions 2 and 3 base pairs within the fragment and at 1 base pair outside the fragment.

Statistical analysis

All statistical analysis were conducted using SciPy (version 1.13) in Python. Linear correlation was measured using the Pearson correlation coefficient. Differences in X were calculated using the Mann-Whitney U test. All reported P values are two-sided.

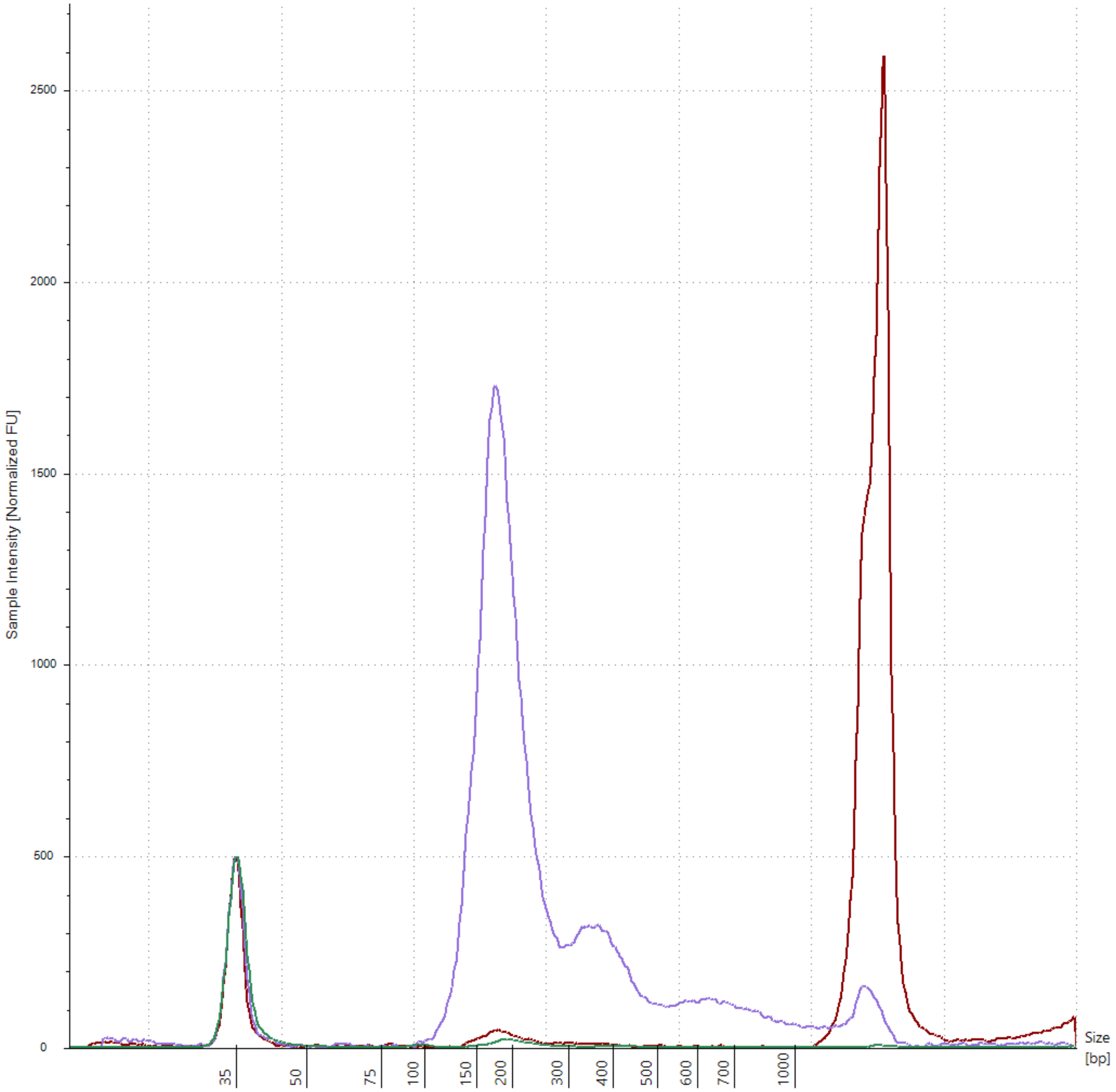


**Fig. S1.**

Electropherogram of DNA isolated from representative samples of plasma from blood tubes (purple line), psDBS (green line), and cDBS (maroon line). psDBS and cDBS samples chosen here showed the highest concentration amongst this sample set.

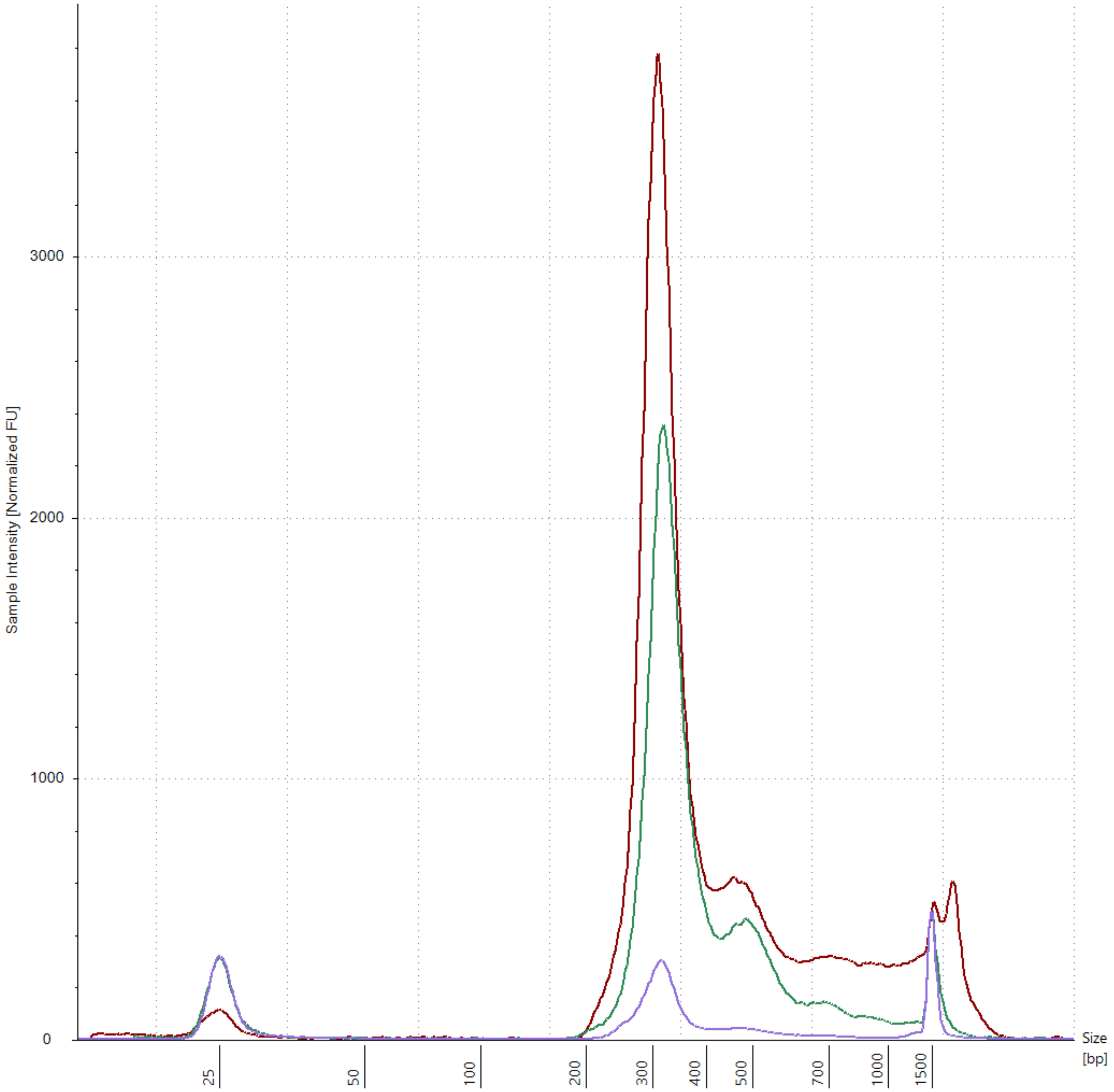


**Fig. S2.**
Electropherogram of whole genome libraries generated from representative samples of plasma from blood tubes (purple line), psDBS (green line), and cDBS (maroon line). Plasma library was diluted 1:10 prior to analysis.

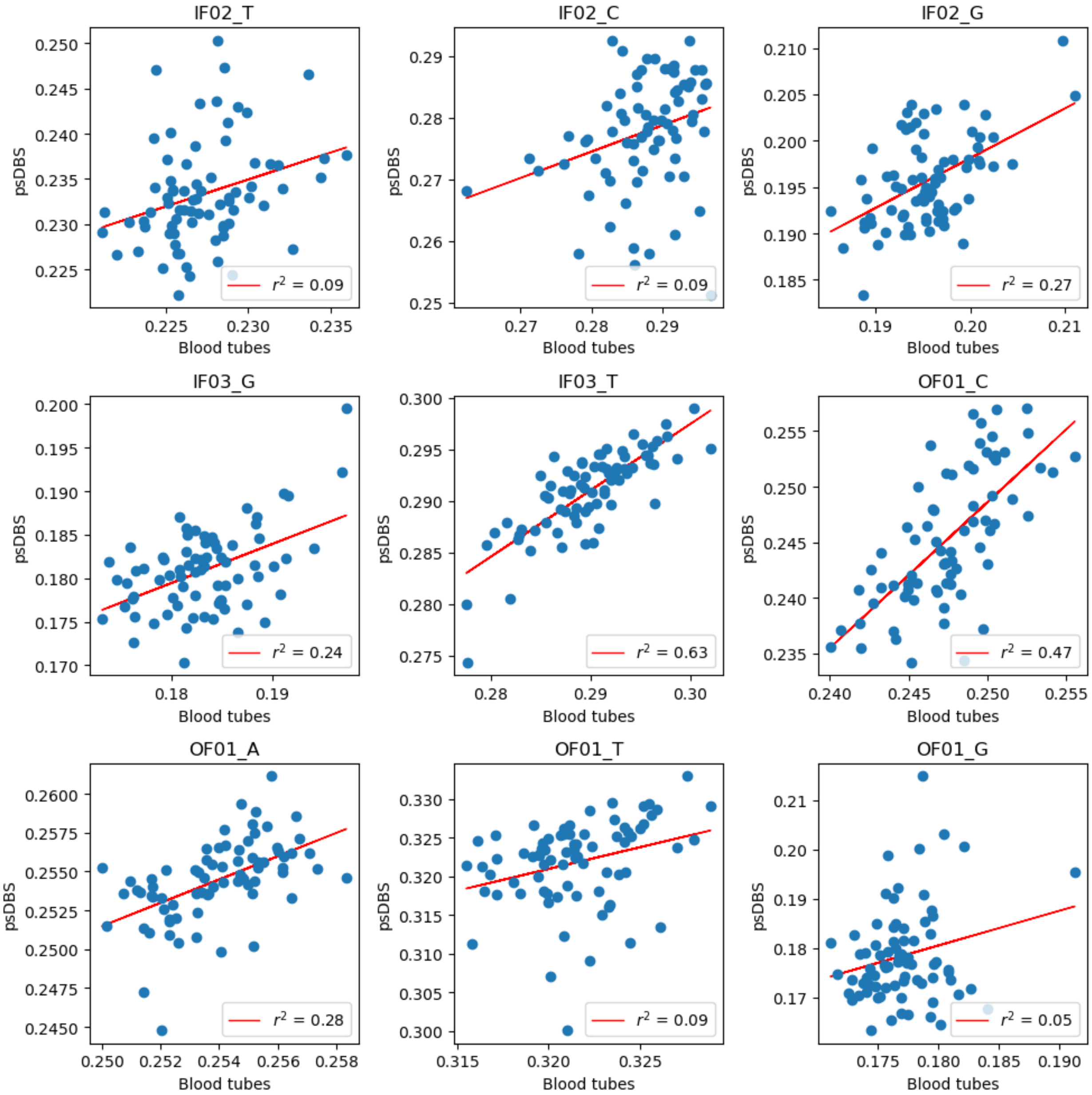


**Fig. S3.**

Correlation in individual nucleotide frequencies surrounding the 5’ fragment end between plasma-separating DBS samples (y-axis) and plasma samples (x-axis). Each plot represents frequency of a single nucleotide at that position relative to the 5’ end. OF01 represents the first position outside the fragment. IF02 represents the second position inside the fragment. IF03 represents the third position inside the fragment.

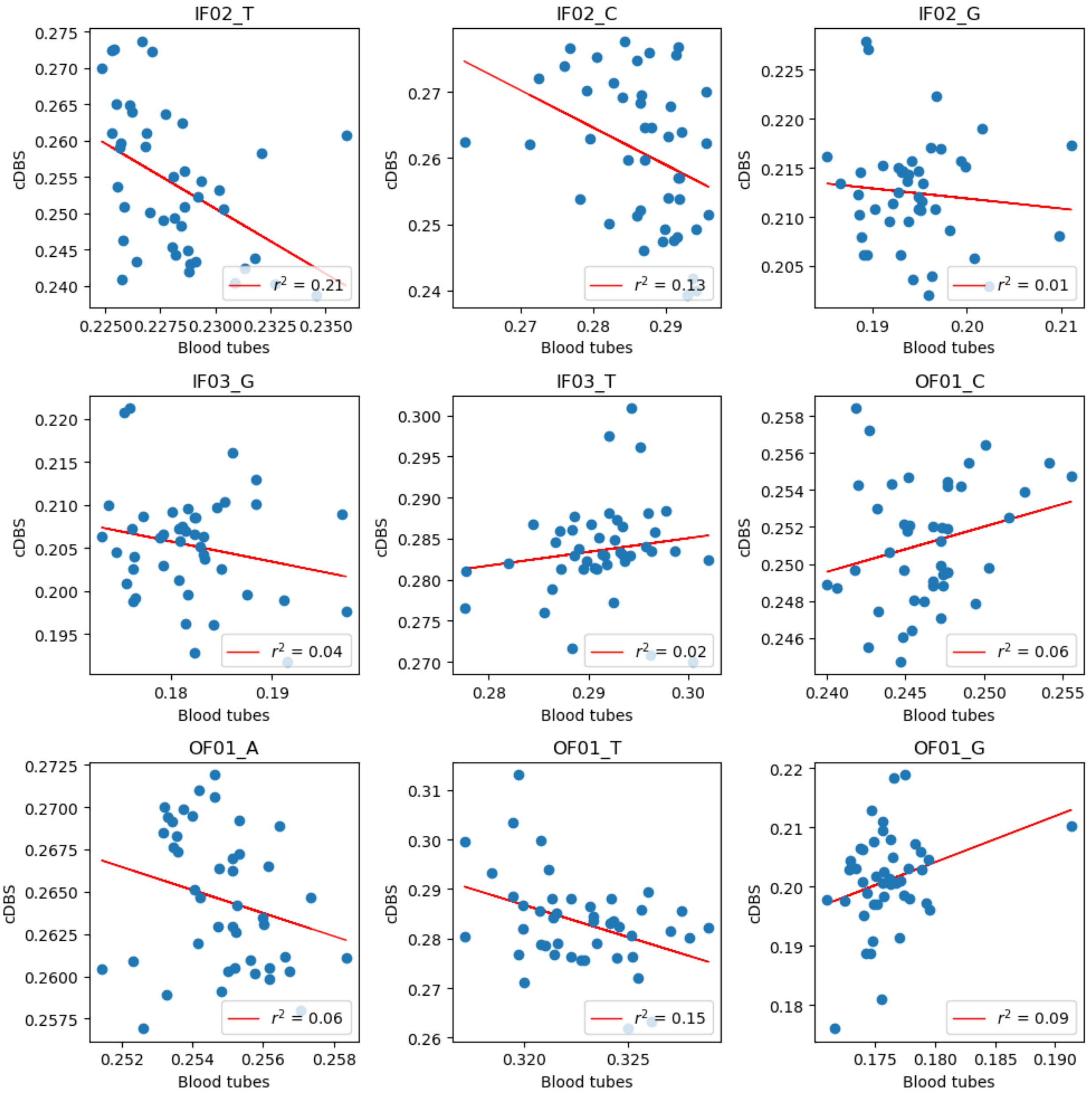


**Fig. S4.**

Correlation in individual nucleotide frequencies surrounding the 5' fragment end between conventional DBS samples (y-axis) and plasma samples (x-axis). Each plot represents frequency of a single nucleotide at that position relative to the 5' end. OF01 represents the first position outside the fragment. IF02 represents the second position inside the fragment. IF03 represents the third position inside the fragment.

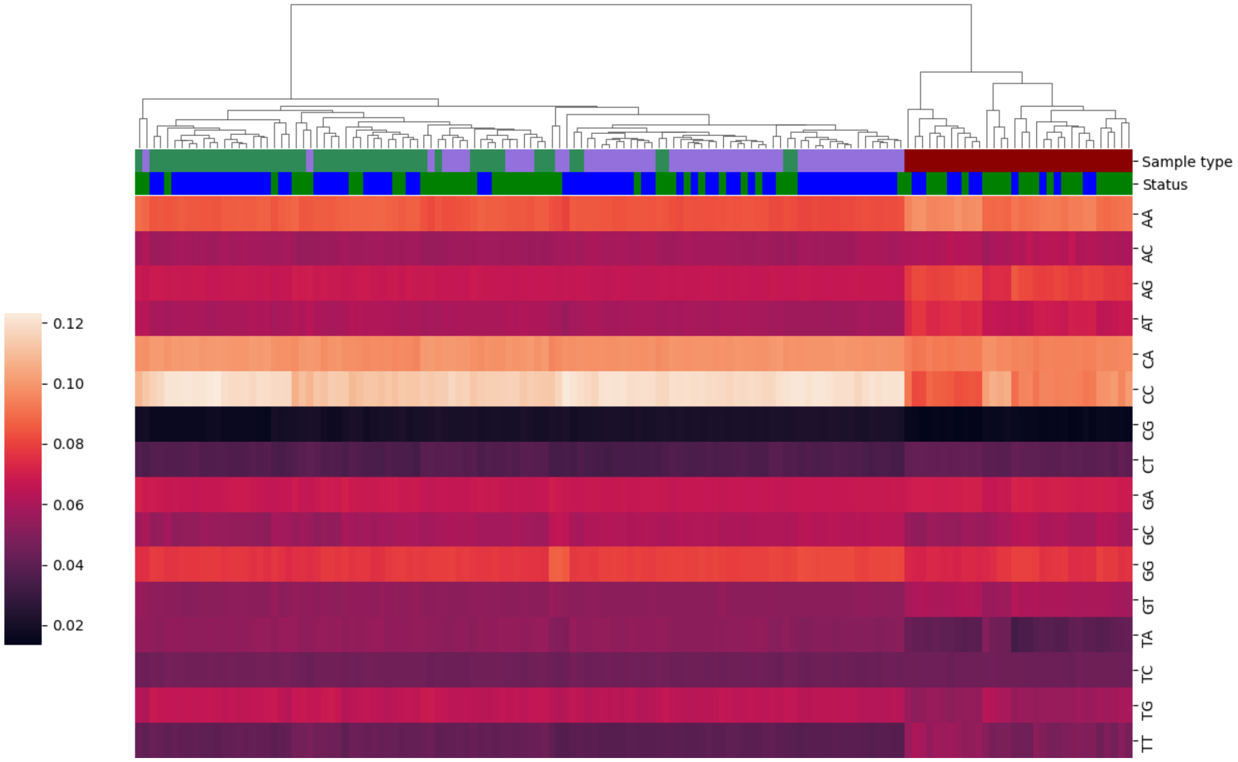


**Fig. S5.**

Hierarchical clustering based on dinucleotide frequencies observed at 5’ ends of cell-free DNA fragments. Plasma samples (purple) and psDBS (green) clustered together compared to cDBS (maroon). The most common dinucleotide observed in plasma samples and psDBS is CC, consistent with prior reports in literature.

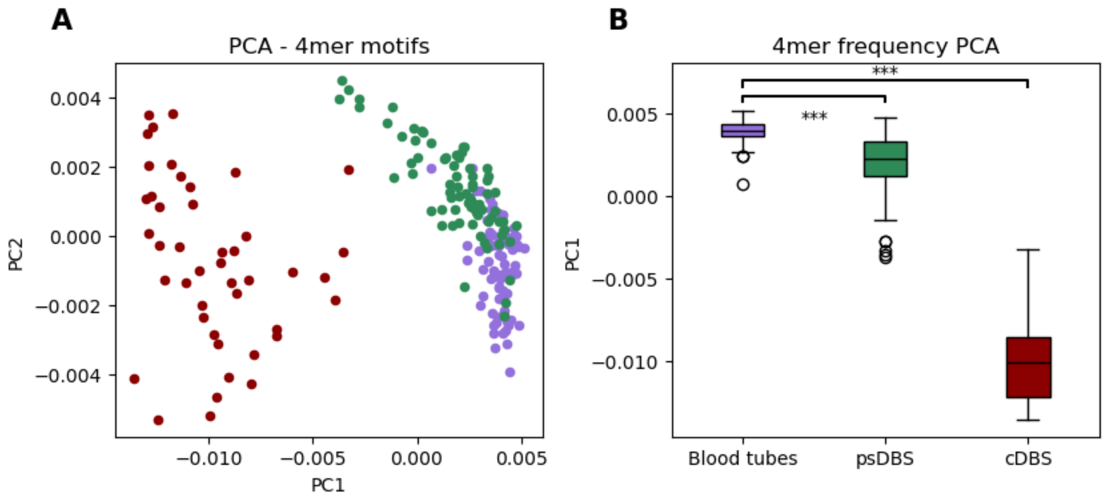


**Fig. S6.**

Comparison of 4-mer frequencies observed at 5' ends of cell-free DNA fragments. **A.** Principal component analysis for dinucleotide frequencies from 5' fragment ends. **B.** Box plot comparing principal component 1 for dinucleotide frequencies from 5' fragment ends across all three sample types.

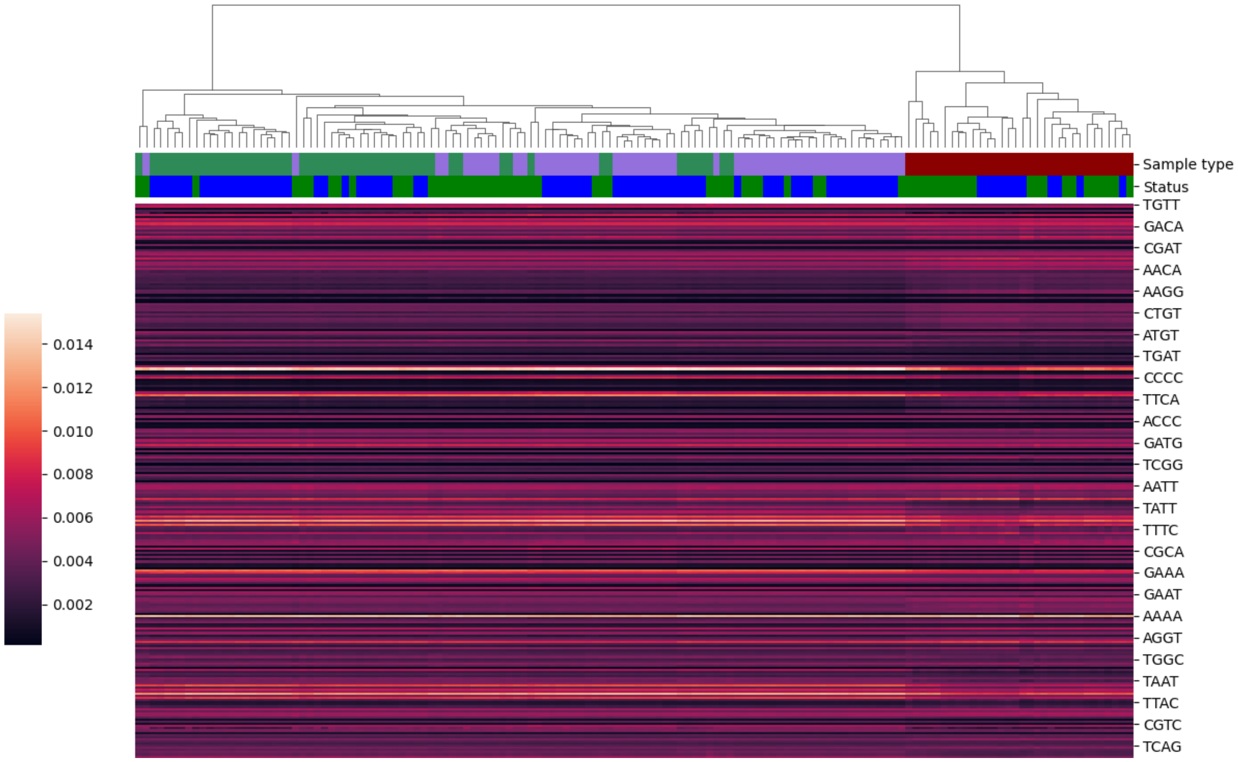


**Fig. S7.**

Hierarchical clustering based on 4-mer frequencies observed at 5' ends of cell-free DNA fragments. Plasma samples (purple) and psDBS (green) clustered together compared to cDBS (maroon).

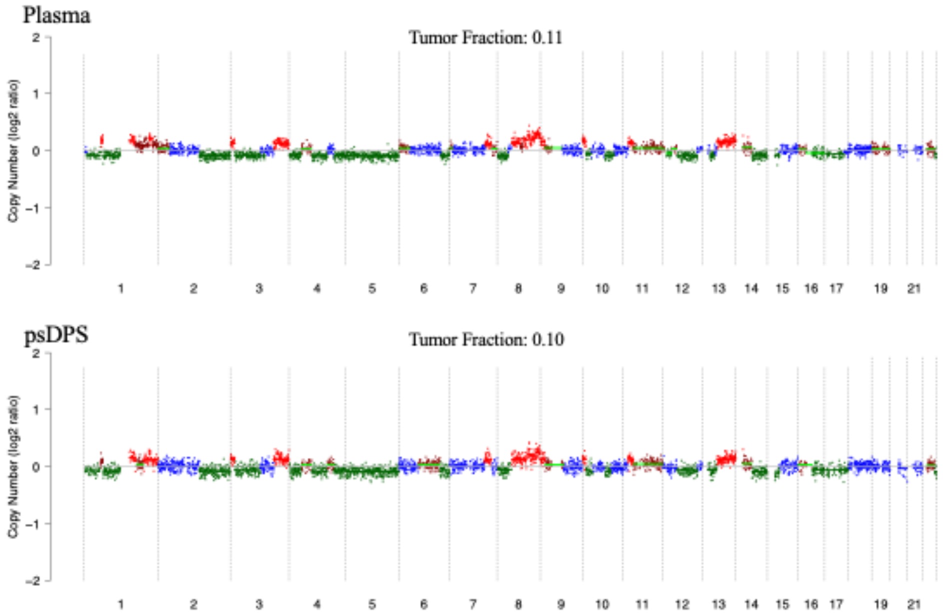


**Fig. S8.**

Comparison of copy-number aberrations and inferred tumor fraction between matched plasma (top) and psDBS (bottom) samples from a patient with Stage IV breast cancer.

**Table S1.**
Demographic characteristics of healthy individuals and patients included in this study.

| Characteristic | | Healthy individuals n (%) | Patients with nonmalignant disease n (%) | Patients with cancer n (%) |
|---|---|---|---|---|
| Total individuals | | 45 (100) | 5 (100) | 25 (100) |
| Age | | | | |
| | 24 and under | 1 (2.2) | 0 (0) | 0 (0) |
| | 25 – 29 | 11 (24.4) | 0 (0) | 0 (0) |
| | 30 – 34 | 2 (4.4) | 0 (0) | 0 (0) |
| | 35 – 39 | 6 (13.3) | 0 (0) | 2 (8) |
| | 40 – 44 | 10 (22.2) | 0 (0) | 0 (0) |
| | 45 – 49 | 1 (2.2) | 0 (0) | 0 (0) |
| | 50 – 54 | 4 (8.9) | 0 (0) | 2 (8) |
| | 55 – 59 | 5 (11.1) | 0 (0) | 6 (24) |
| | 60 and over | 5 (11.1) | 5 (100) | 15 (60) |
| Sex | | | | |
| | Female | 32 (71.1) | 1 (20) | 14 (56) |
| | Male | 13 (28.9) | 4 (80) | 11 (44) |

**Table S2.**
Clinical characteristics of patients with cancer included in this study

| **Characteristic** Tumor type | | **n (%)** |
|---|---|---|
| | Breast | 8 (32) |
| | Endometrium | 4 (16) |
| | Gall bladder | 1 (4) |
| | Head and neck | 5 (20) |
| | Kidney | 2 (8) |
| | Pancreas | 4 (16) |
| | Prostate | 1 (4) |
| Stage | | |
| | I | 9 (36) |
| | II | 5 (20) |
| | III | 7 (28) |
| | IV | 3 (12) |
| | Other | 1 (4) |
